# Dengue Vaccine Effectiveness: Results from a Six-Year Population-Based Cohort Study in Southern Brazil

**DOI:** 10.1101/2023.12.28.23300598

**Authors:** Karin Regina Luhm, Silvia Emiko Shimakura, Sonia Mara Raboni, Magda Clara Vieira da Costa-Ribeiro, Fredi Alexander Diaz-Quijano, Angela Maron de Mello, Lineu Roberto da Silva, Marilene da Cruz Magalhães Buffon, Eliane Mara Cesário Pereira Maluf, Gabriel Graeff, Clara Preto, Gustavo Araújo de Almeida, Gabriela Amanda de Sousa, Elias Teixeira Krainski, Allan Arnold Evans, Denise Siqueira de Carvalho

## Abstract

The alarming growth of dengue worldwide and its social and economic impact have demanded more effective responses for its prevention and control. Currently, the first vaccine approved in Brazil for its prevention, Dengvaxia®, was administered to a target population of around 500,000 residents in southern Brazil. This study reports its effectiveness after a six-year follow-up period from August 2016 to July 2022. Dengue vaccination campaign was carried out in a target population of individuals aged 15–27 in 28 municipalities and 9–44 years in the other two. In this population-based cohort study, exposure to the vaccine included groups with different numbers of doses and adherence to the complete schedule. The primary outcome was probable dengue case. Other endpoints included laboratory-confirmed dengue, serotype, dengue with warning signs or severe illness, and hospitalization. Approximately 60.4% of the participants received at least one vaccine dose. A total of 50,658 probable dengue cases (PDC) were notified of which 15,131 were laboratory-confirmed dengue cases. Overall effectiveness for at least one dose was 33.7% (95% CI: 32.5–34.9) for PDC and 20.1% (95% confidence interval [CI]: 17.1–22.9) for laboratory-confirmed cases. Greater vaccine effectiveness was observed in older individuals and for the DENV-4 and DENV-1 serotypes. The vaccine showed no effectiveness against DENV-2; no DENV-3 cases occurred. A lower hospitalization rate was observed among the vaccinated group. Differences in the incidence of severe dengue cases and warning signs could not draw a definite conclusion. Vaccination was associated with a one-third reduction in the incidence of probable dengue cases. By serotypes, the reduction was significant only for DENV-1 and DENV-4.

**Author Summaries:** Outcomes resulting from dengue mass vaccination remains limited.

Study on Dengvaxia®’s effectiveness in a target population of 501,000 with unknown serostatus. Dengue risk reduction was significant for DENV-1 and DENV-4.

## Introduction

Vector-borne diseases, including dengue, cause approximately 17% of infectious diseases worldwide. It is the fastest-spreading viral infection caused by mosquito bites, endangering 3.97 billion people in 128 countries on nearly every continent, with approximately 400 million infections annually [1]. Incidence rates are highest in South Asia, followed by Southeast Asia, the Caribbean, tropical Latin America, and Central Latin America [2]. In 2022, the Americas reported 2.8 million dengue cases, of which 82% were from Brazil [3]. In the same year, Paraná was Brazil’s third most affected state, with an incidence rate of 1,413.6 per 100,000 inhabitants [4].

There is no specific treatment for dengue, and control strategies for its main vector, *Aedes aegypti*, have presented some limitations [5]. Thus, a vaccine is needed as part of an integrated approach to prevent and control dengue. The first licensed dengue vaccine, Dengvaxia^®^, developed by Sanofi, was introduced in Brazil in December 2015 for individuals aged 9–44 years [6] as a three-dose regimen vaccine with six-month intervals between doses. In clinical trials involving over 30,000 subjects, its overall efficacy against virologically confirmed symptomatic dengue was 65.6% for participants of 9 years of age or older after a 25-month follow-up period. Its efficacy also varied with respect to serotype (DENV-1: 58.4%; DENV-2: 47.1%; DENV-3: 73.6%; DENV-4: 83.2%) [7–9].

This dengue vaccine has been approved in over 20 countries. However, large-scale vaccination campaigns were only organized in Brazil (Paraná state) and the Philippines [6]. The Paraná State Department of Health (SESA/PR) vaccinated free of charge over 300,000 people in 30 municipalities highly affected by dengue between 2016 and 2018, regardless of previous serological status [10], and its effect has been assessed [11]. Hence, we assessed the dengue vaccine’s effectiveness in a population-based cohort six years following the launch of the vaccination initiative in Paraná, Brazil.

## Methods

### Study design and participants

This six-year population-based cohort study focused on a dengue-vaccinated target population residing in 30 municipalities within Paraná state, Brazil, spanning from August 2016 to July 2022.

Paraná is in Brazil’s southern region and dengue vaccination was carried out in the municipalities chosen based on epidemiological criteria. The target population in 28 municipalities comprised individuals aged 15–27 and 9–44 years in other two. The vaccination roll-out consisted of five consecutive phases (Table 1) from August 13, 2016, to December 20, 2018, with intervals of approximately six months [10].

**Table 1.** Dengue vaccination timeline in Paraná, Brazil from 2016 to 2018.

| Phase | Doses administered | Vaccination period |
| --- | --- | --- |
| 1<sup>st</sup> | 1<sup>st</sup> dose | August 13 to September 24, 2016 |
| 2<sup>nd</sup> | 1<sup>st</sup> dose / 2<sup>nd</sup> dose | March 1 to April 7, 2017 |
| 3<sup>rd</sup> | 2<sup>nd</sup> dose / 3<sup>rd</sup> dose | September 20 to November 11, 2017 |
| 4<sup>th</sup> | 2<sup>nd</sup> dose / 3<sup>rd</sup> dose | March 20 to June 29, 2018 |
| 5<sup>th</sup> | 3<sup>rd</sup> dose | November 5 to December 20, 2018 |

Source: SESA/PR

Dengue is a mandatory notification disease, and suspected and confirmed cases nationwide are registered in the National Reportable Disease Information System (Sinan) [12]. Since 1995, dengue has been reported in Paraná, and in recent years, the disease incidence has been the highest ever observed, mainly between 2015–2016 and 2019-2020. Despite this, there is no data on the seroprevalence of the disease in this population. Dengue cases in the 30 municipalities showed a similar pattern to that observed at the state level, with these municipalities accounting for 47.6% of all dengue cases between 2008 and 2016. Four serotypes have circulated in the region since 1995, and DENV-1 has been the most prevalent serotype, except for 2019-2020, in which DENV-2 surpassed the values previously observed for serotype 1 (Figure 1).

**Figure 1.**
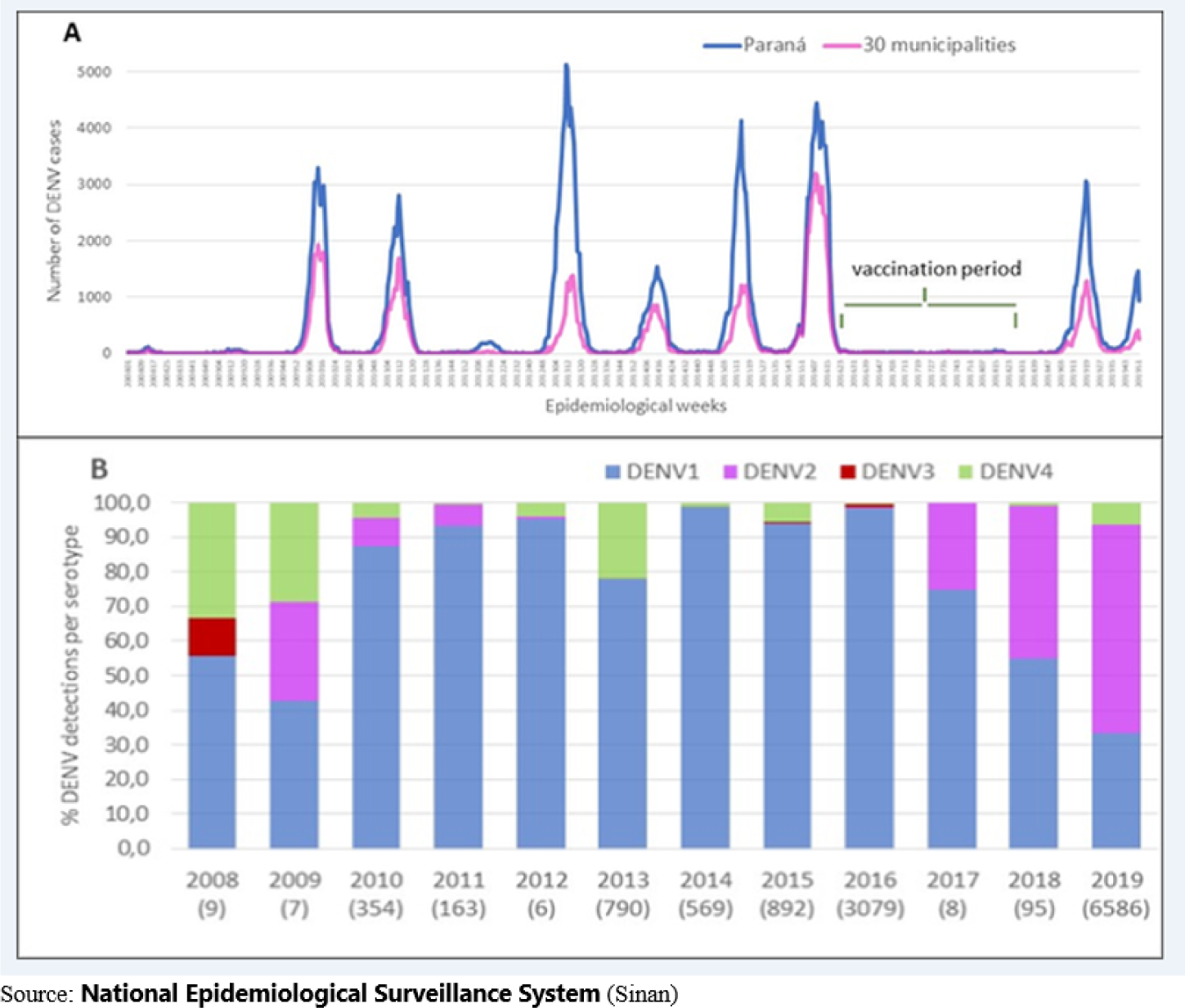
Distribution of probable dengue cases in the state of Paraná and the 30 selected municipalities based on the epidemiological weeks (A), and proportion of dengue serotypes identified in the state of Paraná, Brazil (B), between 2008 and 2019.

The study population was derived from the population defined by SESA/PR to receive public vaccination and included about 500,000 individuals [10]. These individuals were divided into the vaccinated group, constituted of those who had received any number of dengue vaccine doses, and the non-vaccinated group, comprised of individuals who were eligible for vaccination but were not listed in the vaccinated database. The reference population was obtained by subtracting the number of vaccinated individuals from the target population size.

### Procedures

Exposure variables (vaccinations) were obtained from the SESA/PR vaccine database. They were defined based on the number of vaccine doses received (one, two, or three). A subset of individuals who received three doses in consecutive phases, with an interval of ≥ 160 days between doses, was considered to have the complete regimen as recommended and was called the compliance group.

The primary outcome was probable dengue cases (PDC) constituted either by clinical-epidemiological criteria and/or laboratory tests. Secondary outcomes were defined as: i) laboratory- confirmed dengue; ii) confirmed cases with identified serotypes (DENV-1, 2, 3, and 4); iii) cases with warning signs or severe disease identified based on PDC; and iv) dengue hospitalizations with warning signs or severe disease identified based on PDC.

Control variables included sex, age group, and municipality of residence, which may be associated with adherence to the vaccination schedule and disease risk. Age groups were categorized into 9–14, 15-27, and 28–44 years according to the following criteria: a) individuals aged 15–27 years constituted the target population for vaccination in all 30 municipalities; b) individuals aged 9–14 and 28–44 years constituted the target population for vaccination in only two municipalities.

To identify outcomes in vaccinated patients, the Sinan probabilistic record linkage (PRL) procedure was performed in the SESA/PR vaccination database using OpenRecLink 3.1 [14]. The PRL, with an accuracy of 94.1% (95% CI: 91.9–95.7%) and a Kappa concordance index of 96.1% (95% CI: 95.4–96.8%), was validated in a previous study (unpublished).

According to Table 2, we provided different start times for each exposure variable to define follow-up time in the cohort. Among the vaccinated individuals, follow-up was initiated 30 days after the first vaccine dose was administered (Table 2). Among the non-vaccinated individuals, follow-up was initiated 30 days after the end of the vaccination phase, when approximately 75% of vaccinated individuals had received their respective doses.

**Table 2.** Initial follow-up date according to the exposure variables for the vaccinated and non-vaccinated groups.

| Exposure variable | Group |  |
| --- | --- | --- |
|  | Vaccinated | Non-vaccinated |
| **Any number of doses or a single dose** | 30th day after the first vaccine dose | October 23, 2016 (30 days after the end of the first phase of vaccination) |
| **Two doses** | 30th day after the second vaccine dose | May 7, 2017 (30 days after the end of the second phase of vaccination). |
| **Three doses or *compliant*** | 30th day after the third vaccine dose | December 11, 2017 (30 days after the end of the third phase of vaccination) |

For analytical purposes, the follow-up duration for each participant was extended up to the date of their first confirmed dengue episode in case of disease occurrence. For those who remained dengue-free, the predetermined endpoint for follow-up was set at July 31, 2022, with no recorded losses considered in this group.

### Case definitions

Dengue is a mandatorily reportable disease, and the system relies on the notification of all suspected cases of the disease at public and private health facilities based on the initial clinical diagnosis (not laboratory confirmed) [12,15]. The primary outcome was a probable case of dengue (PDC), which includes clinical-epidemiological criteria or laboratory confirmation [13,15], as it is a more sensitive criterion for the diagnosis of dengue and is helpful in the context of epidemiological surveillance of the disease. Laboratory-confirmed dengue was defined as PCR, NS1, or IgM positive test. IgM result was only considered if performed 90 days after any vaccine dose for those vaccinated. *Statistical analysis*

Incidence rates per 10,000 person-years (IR/10^4^py) and incidence rate ratios (IRR) were calculated for both primary outcomes under consideration. These measures were determined for individuals who received at least one, two, or three vaccine doses or complete regimen as recommended compared to non-vaccinated individuals. Other secondary outcomes were evaluated in patients who received at least one dose.

Poisson regression was used to adjust for confounders, with the time of follow-up used as an offset. Age, sex, and municipality were included in the models. Individuals aged 9–14 and 28–44 years (11.5%) were excluded, except when specific analysis by age group was performed, as they were restricted to only two municipalities.

Vaccine effectiveness and 95% confidence intervals (95% CIs) were calculated using the formula:

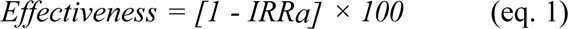

where IRRa is the adjusted incidence rate ratio. Statistical analyses were performed using R 2020 (version 4.0.2) and Stata® (version 15.1) [16,17]. Absolute risk reduction (ARR) and the number necessary for vaccine (NNV) were also calculated and presented for each exposure regimen [18].

For sensitivity analysis, the effectiveness of PDC was evaluated considering the period from 2019 onwards.

This study was approved by the Research Ethics Committee of the Federal University of Paraná (N# 2,308,662) and registered in the institution’s Integrated Agreement Management System under Process number 23075.040144/2019-49 and published in the DOU on 04/June/ 2020.

## Results

This cohort consisted of 501,208 individuals from the vaccination’s target population program, 302,603 (60.4%) individuals who received at least one vaccine dose and 198,605 (39.6%) unvaccinated individuals. A total of 142,687 individuals received the three planned doses, among which 122,564 met the compliance criteria.

Demographic characteristics and vaccination-related aspects are shown in Table 3. A higher percentage of men received at least one vaccine dose (51.1%) and women had a higher compliance rate (52.4%). The highest frequency of dengue notifications was recorded in 2020 (44.9%).

**Table 3.** Demographic characteristics, exposure, and dengue notifications in the target population in the 30 municipalities in Paraná from epidemiological week 33/2016 to 52/2019.

| Characteristic | Vaccinated |  |  |  | Non-vaccinated |  | Total |  |
| --- | --- | --- | --- | --- | --- | --- | --- | --- |
|  | *Compliant* |  | Total vaccinated |  | (Total: 198,605) |  | (Total: 501,208) |  |
|  | (Total: 122,614) |  | (Total: 302,603) |  |  |  |  |  |
|  | n | % | n | % | n | % | n | % |
| **Sex** |  |  |  |  |  |  |  |  |
| Male | 59,914 | 48.9 | 158,654 | 52.4 | 90,391 | 45.5 | 249,045 | 49.7 |
| Female | 62,700 | 51.1 | 143,949 | 47.6 | 108,214 | 54.5 | 252,163 | 50.3 |
| **Age group** |  |  |  |  |  |  |  |  |
| 9 to 14* | 7,980 | 6.5 | 14,409 | 4.8 | 4,894 | 2.5 | 19,303 | 3.9 |
| 15 to 19 | 48,718 | 39.7 | 111,993 | 37.0 | 56,354 | 28.4 | 168,347 | 33.6 |
| 20 to 24 | 32,776 | 26.7 | 94,124 | 31.1 | 79,822 | 40.2 | 173,946 | 34.7 |
| 25 to 27 | 16,813 | 13.7 | 48,509 | 16.0 | 52,811 | 26.6 | 101,320 | 20.2 |
| 28 to 44* | 16,327 | 13.3 | 33,568 | 11.1 | 4,724 | 2.4 | 38,292 | 7.6 |
| **Cohort group** |  |  |  |  |  |  |  |  |
| First | 104,484 | 85.2 | 205,927 | 68.1 | 198,605 | 100.0 | 404,532 | 80.7 |
| Second | 18,130 | 14.8 | 96,676 | 31.9 | .. | .. | 96,676 | 19.3 |
| **Number of doses** |  |  |  |  |  |  |  |  |
| 3 doses | 122,614 | 100.0 | 142,700 | 47.2 | .. | .. | 142,700 | 28.5 |
| 2 doses | 0 | .. | 78,337 | 25.9 | .. | .. | 78,337 | 15.6 |
| 1 dose | 0 | .. | 81,566 | 27.0 | .. | .. | 81,566 | 16.3 |
| None | 0 | .. | 0 | .. | 198,605 | 100.0 | 198,605 | 39.6 |
| **Notifications** |  |  |  |  |  |  |  |  |
| 2016 | 349 | 7.9 | 935 | 8.4 | 1,029 | 7.8 | 1,964 | 8.1 |
| 2017 | 831 | 18.7 | 2,280 | 20.6 | 2,253 | 17.1 | 4,533 | 18.7 |
| 2018 | 533 | 12.0 | 1,281 | 11.6 | 1,584 | 12.0 | 2,865 | 11.8 |
| 2019 | 2,729 | 61.4 | 6,591 | 59.4 | 8,300 | 63.0 | 14,891 | 61.4 |
| **Total** | 4,442 | 100.0 | 11,087 | 100.0 | 13,166 | 100.0 | 24,253 | 100.0 |

The denominator of the proportions was the total number of individuals in each group (compliant, vaccinated, non-vaccinated, and total cohort). Total vaccinated includes individuals who received any number of vaccine doses. \* For the municipalities of Assaí and Paranaguá, the study population was only composed of the age groups 9–14 and 28–44 years.

Among the 90,373 notifications recorded from 2016–2022, PDC numbered 50,658, of which 24,219 (47.8%) in the vaccinated group (Table 4). A total of 15,131 (16.7%) were laboratory-confirmed cases with 8,364 (55.3%) in the vaccinated group. Warning signs were identified in 912 cases, with 520 (57.0%) in the vaccinated group.

**Table 4.**
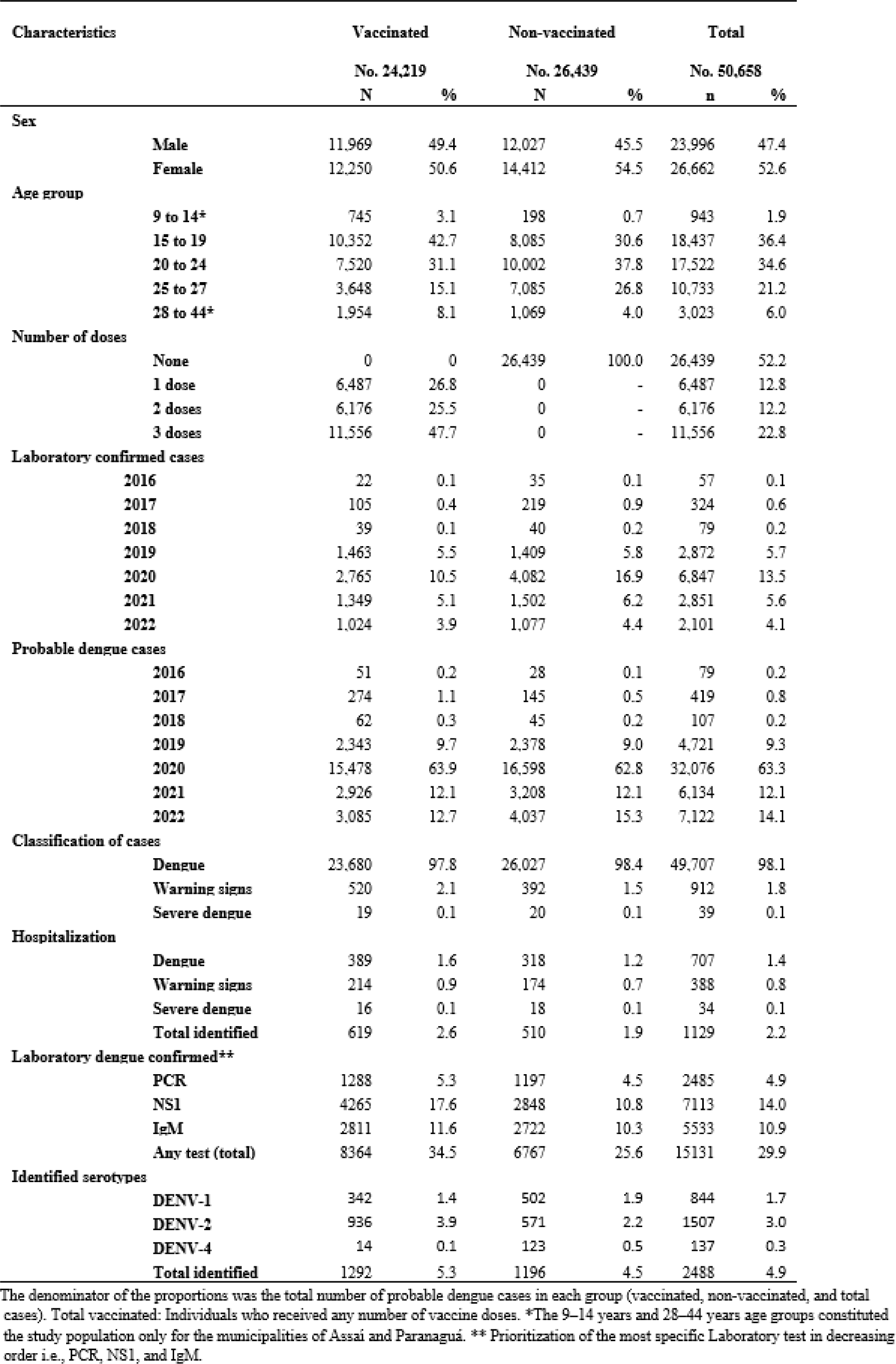
Probable dengue cases (PDC) in the target population in 30 municipalities in Paraná.

Thirty-nine severe cases were observed among PDC, 19 in the vaccinated group and 20 in the non-vaccinated. Dengue-related deaths were reported in both groups during the study period, 5 cases in the vaccinated and 14 occurrences in the non-vaccinated (IR 0.39; 95% CI: 0.14-1.08). Of the 1,129 hospitalizations, 619 (54.0%) were in the vaccinated group, of which 16 (2.6%) involved severe forms, while of the 510 hospitalizations in the non-vaccinated group, 18 (3,5%) were severe.

Vaccine effectiveness on PDC ranged from 31.5% (95% CI: 29.5–33.5) to 35.1% (95% CI: 33.3– 36.8) among those with two doses and those with compliance, respectively (Table 5).

**Table 5.**
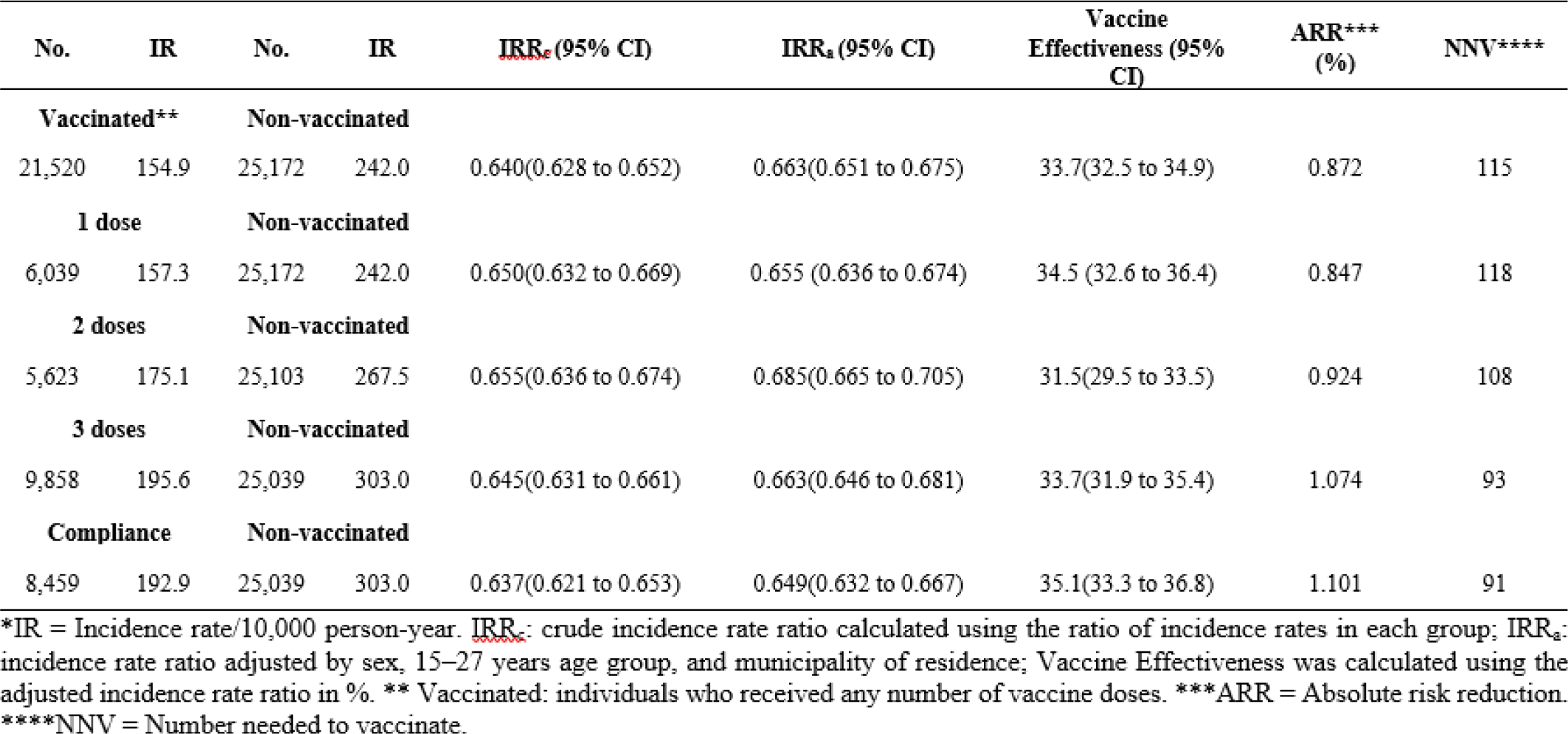
Number of probable dengue cases, incidence rates in 10, 000 person-year, crude and adjusted incidence rate rations, vaccine effectiveness, absolute risk reduction and number needed to vaccinate according to exposure characteristics and primary outcome in 30 municipalities in Paraná.

| No. | IR | No. | IR | IRR<sub>c</sub> (95% CI) | IRR<sub>a</sub> (95% CI) | Vaccine Effectiveness (95% CI) | ARR*** (%) | NNV**** |
| --- | --- | --- | --- | --- | --- | --- | --- | --- |
| **Vaccinated\*\*** |  | **Non-vaccinated** |  |  |  |  |  |  |
| 21,520 | 154.9 | 25,172 | 242.0 | 0.640(0.628 to 0.652) | 0.663(0.651 to 0.675) | 33.7(32.5 to 34.9) | 0.872 | 115 |
| **1 dose** |  | **Non-vaccinated** |  |  |  |  |  |  |
| 6,039 | 157.3 | 25,172 | 242.0 | 0.650(0.632 to 0.669) | 0.655 (0.636 to 0.674) | 34.5 (32.6 to 36.4) | 0.847 | 118 |
| **2 doses** |  | **Non-vaccinated** |  |  |  |  |  |  |
| 5,623 | 175.1 | 25,103 | 267.5 | 0.655(0.636 to 0.674) | 0.685(0.665 to 0.705) | 31.5(29.5 to 33.5) | 0.924 | 108 |
| **3 doses** |  | **Non-vaccinated** |  |  |  |  |  |  |
| 9,858 | 195.6 | 25,039 | 303.0 | 0.645(0.631 to 0.661) | 0.663(0.646 to 0.681) | 33.7(31.9 to 35.4) | 1.074 | 93 |
| **Compliance** |  | **Non-vaccinated** |  |  |  |  |  |  |
| 8,459 | 192.9 | 25,039 | 303.0 | 0.637(0.621 to 0.653) | 0.649(0.632 to 0.667) | 35.1(33.3 to 36.8) | 1.101 | 91 |

\*IR = Incidence rate/10,000 person-year. IRR<sub>c</sub>: crude incidence rate ratio calculated using the ratio of incidence rates in each group; IRR<sub>a</sub>: incidence rate ratio adjusted by sex, 15–27 years age group, and municipality of residence; Vaccine Effectiveness was calculated using the adjusted incidence rate ratio in %. \*\* Vaccinated: individuals who received any number of vaccine doses. \*\*\*ARR = Absolute risk reduction. \*\*\*\*NNV = Number needed to vaccinate.

Based on laboratory- confirmed cases, overall effectiveness was 20.1 (95% CI: 17.1-22.9) for at least one dose (Table 6). It ranged from 17.5 (95% CI:12,9–21.9) among those who received two doses to 24.3% (95% CI: 20.1–28.2) for those with one dose.

**Table 6.** Number of laboratory confirmed dengue cases, incidence rates in 10, 000 person-year, crude and adjusted incidence rate rations, vaccine effectiveness, absolute risk reduction and number needed to vaccinate according to exposure characteristics and primary outcome in 30 municipalities in Paraná.

| No. | IR | No. | IR* | IRR<sub>c</sub> (95% CI) | IRR<sub>a</sub> (95% CI) | Vaccine Effectiveness (95% CI) | ARR*** (%) | NNV**** |
| --- | --- | --- | --- | --- | --- | --- | --- | --- |
| **Vaccinated\*\*** |  | **Non-vaccinated** |  |  |  |  |  |  |
| 6,869 | 51.4 | 6,074 | 62.8 | 0.818(0.790 to 0.847) | 0.799 (0.771 to 0.829) | 20.1(17.1 to 22.9) | 0.142 | 876 |
| **1 dose** |  | **Non-vaccinated** |  |  |  |  |  |  |
| 1,817 | 49.2 | 6,074 | 62.8 | 0.783(0.743 to 0.826) | 0.757 (0.718 to 0.799) | 24.3 (20.1 to 28.2) | 0.136 | 735 |
| **2 doses** |  | **Non-vaccinated** |  |  |  |  |  |  |
| 1,810 | 58.3 | 6,018 | 68.7 | 0.849(0.805 to 0.895) | 0.825(0.781 to 0.871) | 17.5(12.9 to 21.9) | 0.104 | 966 |
| **3 doses** |  | **Non-vaccinated** |  |  |  |  |  |  |
| 3,242 | 66.5 | 5,987 | 77.2 | 0.861(0.825 to 0.899) | 0.817(0.777 to 0.858) | 18.3(14.2 to 22.3) | 0.107 | 934 |
| **Compliance** |  | **Non-vaccinated** |  |  |  |  |  |  |
| 2,825 | 66.6 | 5,987 | 77.2 | 0.863(0.825 to 0.903) | 0.806(0.766 to 0.849) | 19.4(15.1 to 23.4) | 0.106 | 945 |

\*IR = Incidence rate/10,000 person-year. IRR<sub>c</sub>: crude incidence rate ratio calculated using the ratio of incidence rates in each group; IRR<sub>a</sub>: incidence rate ratio adjusted by sex, 15–27 years age group, and municipality of residence; Vaccine Effectiveness was calculated using the adjusted incidence rate ratio in %. \*\* Vaccinated: individuals who received any number of vaccine doses. \*\*\*ARR = Absolute risk reduction. \*\*\*\*NNV = Number needed to vaccinate.

Higher effectiveness was observed in the 28–44 age group (78.1%; 95% CI: 76.2–79.9%), irrespective of the number of vaccine doses (Table 7). Data for the 28–44 age group were limited to the municipalities of Paranaguá and Assaí. When vaccine effectiveness was stratified by serotype, it was found to be 51.9% (95% CI: 44.6–58.2) and 92.0% (95% CI: 86.2–95.4) for DENV-1 and DENV-4, respectively. No effectiveness was found for DENV-2 (−10.8%; 95% CI: −23.5–0.60), and no DENV-3 cases were reported. Hospitalization was reduced among the vaccinated group (16.5%; CI: 5.5 to 26.3). Dengue cases with warning signs and/or severe forms presented wide confidence intervals for effectiveness, and no definite conclusions may be drawn.

**Table 7.** Other secondary outcomes, incidence rates in 10, 000 person-year, crude and adjusted incidence rate rations, vaccine effectiveness, absolute risk reduction and number needed to vaccinate according to exposure characteristics in 30 municipalities in Paraná.

|  | No. | IR | No. | IR | <u>IRR<sub>c</sub></u> (95% CI) | IRR<sub>a</sub> (95% CI) | Vaccine Effectiveness (95% CI) | ARR*** (%) | NNT**** |
| --- | --- | --- | --- | --- | --- | --- | --- | --- | --- |
| **Age groups** | **Vaccinated\*\*** |  | **Non-vaccinated** |  |  |  |  |  |  |
| 9 to 14 years | 745 | 91.9 | 198 | 71.1 | 1.292(1.103 to 1.519) | 1.090(0.928 to 1.281) | -9.0(-28.1 to 7.2) | -0.201 | - |
| 15 to 27 years | 21,520 | 154.9 | 25,172 | 242.0 | 0.640(0.628 to 0.652) | 0.672((0.660 to 0.685) | 32.8 (31.5 to 34.0) | 0.872 | 115 |
| 28 to 44 years | 1,954 | 103.4 | 1,069 | 425.6 | 0.243(0.225 to 0.262) | 0.219((0.201 to 0.238) | 78.1 (76.2 to 79.9) | 3.222 | 31 |
| **Serotypes** | **Vaccinated\*\*** |  | **Non-vaccinated** |  |  |  |  |  |  |
| DENV-1 | 333 | 2.5 | 495 | 5.2 | 0.485(0.420 to 0.558) | 0.481(0.418 to 0.554) | 51.9(44.6 to 58.2) | 0.027 | 4 |
| DENV-2 | 836 | 6.4 | 546 | 5.8 | 1.102(0.988 to 1.230) | 1.108(0.994 to 1.235) | -10.8(-23.5 to 0.60) | 0.006 | 17 |
| DENV-4 | 14 | 0.1 | 123 | 1.3 | 0.082(0.044 to 0.143) | 0.080(0.046 to 0.139) | 92.0(86.2 to 95.4) | 0.012 | 8 |
| **Clinical characteristics** | **Vaccinated\*\*** |  | **Non-vaccinated** |  |  |  |  |  |  |
| Alarm/Severe signs | 509 | 3.9 | 405 | 4.3 | 0.905(0.792 to 1.033) | 0.901(0.789 to 1.029) | 9.9(-2.9 to 21.1) | 0.004 | 25 |
| Severe dengue | 18 | 0.14 | 20 | 0.21 | 0.648(0.323 to 1.290) | 0.588(0.304 to 1.137) | 41.2(-13.7 to 69.6) | 0.001 | 133 |
| Hospitalization | 537 | 3.9 | 484 | 4.7 | 0.830(0.733 to 0.941) | 0.835(0.737 to 0.945) | 16.5(5.5 to 26.3) | 0.008 | 13 |

\*IR = Incidence rate/10,000 person-year. IRR<sub>c</sub>: crude incidence rate ratio calculated using the ratio of incidence rates in each group; IRR<sub>a</sub>: incidence rate ratio adjusted by sex and 15–27 years age group; Vaccine Effectiveness was calculated using the adjusted incidence rate ratio in %. \*\* Vaccinated: individuals who received any number of vaccine doses. \*\*\*ARR = Absolute risk reduction. \*\*\*\*NNV = Number needed to vaccinate.

In the sensitivity analysis, 21,252 cases were identified among individuals who received at least one dose (IR 245.2/10^4^py) and 24,993 among non-vaccinated individuals (IR 399.1/10^4^py). The measured vaccine effectiveness was 36.6% (95% CI: 35.4–37.8), similar to that obtained in the primary analysis with PDC.

## Discussion

In this study, we report the findings on the effectiveness of dengue vaccine, administered in a large population-based cohort in 30 municipalities in Paraná, Southern Brazil [10], after six years of monitoring. Effectiveness was higher among vaccinated individuals but lower than that obtained in efficacy studies [9]. There was significant variability in vaccine effectiveness (VE) for the different viral subtypes; VE was highest for DEN-4 and DEN-1 and lowest for DEN-2 (negative VE values). The VE for serotype 3 could not be evaluated. The high prevalence of serotype 2 during the study period may explain the differences in effectiveness observed here and previous clinical trials. PDC was used as the primary endpoint as it better represents the real scenario of health services in diagnosing dengue in countries like Brazil [18].

Since 2014, when Brazil adopted the WHO classification for dengue, PDC has been the parameter used for dengue disease monitoring through epidemiological surveillance [15]. The signs and symptoms of dengue are well known by health professionals in endemic areas, favoring homogeneity in diagnosing the disease among vaccinated and non-vaccinated individuals. Furthermore, it is crucial to work with probable cases to respond to epidemics promptly, as case confirmation might be expensive and time-consuming [18].

The challenge in anti-dengue vaccine effectiveness studies is defining the primary endpoint. Ideally, this should be based on laboratory-confirmed disease. Laboratory confirmation, however, is not feasible in regions where thousands of suspected cases of dengue occur each year. Therefore, surveillance systems conclude reported cases based on clinical-epidemiological criteria and, anti-dengue vaccine efficacy studies should consider this criterion in their analysis of results.

Dengue notification processes were affected by the outbreak of the large-scale Zika virus in 2014, which spread across the Pacific and reached Brazil, causing an explosive epidemic in South America. Zika infection can be misdiagnosed clinically as Dengue fever, reducing the accuracy of the clinical-epidemiological diagnosis. Moreover, the high genetic similarity between both viruses contributes to a substantial cross-reactivity in the antibody responses, compromising the results of point-of-care serological tests [19].

However, evaluating VE using laboratory methods exclusively presents limitations. Their use depends on the availability of infrastructure and laboratory capacity, making it challenging to offer and apply these procedures universally across municipalities and healthcare sites. Testing rates differed between municipalities, although there was no significant difference in laboratory testing between vaccinated and non-vaccinated individuals. According to De Smedt et al. [20], the laboratory criterion may introduce a differential misclassification bias by underestimating effectiveness, as observed by comparing PDC-based effectiveness estimates with PCRNS1 endpoints. Furthermore, adding IgM tests to the PDC for case identification ratified the primary outcome-based evaluation.

The “exposure to at least one dose” criterion seems the most appropriate because it represents the natural condition of community intervention and reflects population adherence to multiple-dose vaccinations [10]. There was no difference between compliant individuals and those who received the three doses. However, further studies are needed to compare regimens with different numbers of doses, considering different epidemiological contexts [21], the prevalence of circulating serotypes [22], and the change in vaccine recommendations concerning previous seropositivity [23-25].

Effectiveness was higher in older age groups. Since there is a greater probability of previous dengue infection in older individuals, these data corroborate the results of efficacy studies [7,9,26,27]. Although the basal seroprevalence of dengue in Paraná is unknown, a survey of dengue seroprevalence in a Brazilian city with a history of recurrent epidemics showed seropositivity values ranging from 63– 75% in the 10–19 and 40–59 years age groups, respectively [28].

In line with the findings of efficacy studies (CYD14 and CYD15) [9], effectiveness varied according to viral serotype, with the highest values observed for DENV-4 and DENV-1. Notably, global vaccine effectiveness is significantly associated with serotype-related efficacy and depends on identified circulating serotypes, baseline serostatus, and dengue seroprevalence during the study period [22].

All four dengue serotypes have been circulating in Paraná since 1995, including areas where the vaccination campaign was carried out. Throughout the epidemic years spanning from 2008 to 2016, DENV-1 predominated, with minimal circulation of DENV-2 and DENV-4, and negligible presence of DENV-3. However, in 2017, there was an important upsurge in the circulation of DENV-2. In 2019- 2020, it was responsible for two-thirds of cases, with a low DENV-1 (37%) and 4 (8%) circulation and the absence of DENV-3 observed. In 2021-2022, there was a rise in the DENV-1 detection, but DENV- 2 was predominant [4].

The ineffectiveness of DENV-2 observed in the study population regardless of the serostatus was previously reported by Sabchareon et al. (2012) [29] in a phase IIb Dengvaxia^®^ efficacy study in Thailand and subsequently in the CYD14 and CYD15 studies [30]. The antigenic incompatibility between the vaccine virus (Asian-American genotype) and DENV-2 circulating in Southeast Asia (Asian genotype 1) and the unbalanced replication of vaccine viruses is likely responsible for the variability in vaccine effectiveness against different viral types. [31]. Furthermore, Heinen et al. studies indicate that, among children who were seronegative for DENV at baseline, impaired replication of DENV-4 from the tetravalent vaccine stimulates antibodies capable of neutralizing DENV-1 and DENV-3 *in vitro* but not protecting *in vivo* [32].

Further, studies have shown that viruses obtained directly from infected patients’ plasma are significantly more infectious than those obtained from cell cultures. Although genotypically identical, DENVs obtained in cell lines are structurally immature and hypersensitive to neutralization by human antibodies compared to DENVs of an infected patient. This phenotypic difference is due to the mature state of the virion obtained in plasma samples compared with immature particles abundantly recovered in cell cultures. In this way, the antibodies induced by the vaccine efficiently neutralize the DENVs in cell culture assays, but they are ineffective in protecting the vaccinated individual [33].

Tetravalent dengue vaccines have been developed on the premise that intra-serotype variability does not interfere with vaccine-generated immune responses. However, in a recent analysis of the genetic diversity of DENV-2 pre-membrane (Pre-M) and envelope (E) proteins, its genotypic variants were evaluated, and were found to be neutralized in different ways by monoclonal antibodies and polyclonal sera derived from DENV-2-infected individuals. They also showed differential responses to antibodies induced by both monovalent dengue 2 vaccines and tetravalent vaccines [34]. Therefore, a high antigenic match between vaccine strains and circulating DENVs may be essential to achieve high vaccine efficacy [30].

A study conducted in Brazil using clinical samples from two cities of São Paulo in 2019, a new genotype, DENV-2 III BR4, was identified. It was probably introduced in Brazil in 2014 and has replaced other DENV-2 strains circulating in the region [35]. However, in this study, we did not perform a molecular analysis of DENV-2 strains circulating in the studied municipalities. Dengue viruses are constantly under selective pressure, with the consequent emergence of new genotypes that progressively replace previous strains. Thus, the possibility of DENV immune response evasion should be considered in designing vaccines against dengue, which will probably systematically include viral genotypes circulating in the region at that time [22, 29–31, 34]. Therefore, viral genetic surveillance studies are required to understand better the relationship between the vaccine, viral evolution, and dengue virus genotypic variability.

Vaccine effectiveness was not statistically significant in cases with warning signs, although this outcome was not evaluated in efficacy studies. Few severe cases or warning signs resulted in low accuracy, limiting the assessment of these outcomes. However, it is essential to note that higher incidence rates of hospitalization due to severe cases and warning signs occurred in the non- vaccinated group. The increased risk of hospitalization observed among seronegative participants in follow-up studies led to recommending the vaccine only to seropositive individuals. [24,25]. The results obtained in this study refer to a population with no information about their previous disease situation. Despite this, hospitalization in dengue cases with alarming or severe signs did not pose a greater risk.

The present study was based on a mixed population of dengue-naïve and previously infected individuals when the vaccine was still indicated for all living in regions with high endemicity. From 2018 onwards, the vaccine has only been indicated for people with previous dengue infection (i.e., seropositive) [21]. Therefore, the replication of this study is precluded. Considering future scenarios of effectiveness evaluation, given that the vaccine’s efficacy is increased in baseline seropositive subjects the results of studies limited to seropositive populations would potentially be improved compared to this present study.

This study was conducted using secondary databases, which have some limitations. The probabilistic matching of three bases minimized these limitations, allowing us to retrieve individual information with high agreement and accuracy, exclude duplicates, and complement data. Owing to the data sources used, we could not individually assess the socioeconomic and educational characteristics of the target population, which may be associated with vaccine adherence, health care, and the risk of dengue infection. Studies on factors affecting the acceptance of the dengue vaccine have reported controversial data regarding the effects of socioeconomic status [36,37] and schooling [36–39]. Previous experiences with dengue may also constitute confounding factors, which lead to the vaccination of individuals at a higher risk of developing dengue [37]; however, other studies have not found this association [36,39]. In our study, conditioning for a municipality variable, age, sex, and other characteristics of the municipality are controlled, as they are associated with both the exposure variable (vaccine) and the outcome (notification), minimizing these biases.

The vaccine has proven effective in reducing the incidence of symptomatic dengue in clinical trials, but evidence is lacking regarding the impact it would have on epidemiological surveillance. In this study, we estimated a reduction in reported PDC, representing 33.7% among individuals who received at least one dose of the vaccine.

DENVs are heterogeneous and dynamic, with consequent implications for vaccine research. Recent human natural dengue infection studies have shown that only the presence of anti-DENV antibodies is not enough to define neutralization and protection, as not all neutralizing antibodies are equally protective to the same degree. Therefore, the quality and quantity of anti-DENV neutralizing antibodies are critical. Most importantly, dengue research must focus on understanding DENV immunity and searching for reliable protection markers [40].

The evaluation of effectiveness on laboratory-confirmed cases and serotype-specific infections would have the limitation that the identification of the outcome would be conditioned to a series of known and unknown factors that affect access to diagnosis. These factors can either be a consequence of consultation patterns, also associated with vaccination adherence, or be associated with immunization outcomes (manifestations and disease severity). Thus, there would be a high risk of selection bias when focusing the analysis only on confirmed ones [37]. By estimating the impact of vaccination on the total PDC, which more sensitively and robustly represents the burden of the disease in the health system, these results become more relevant for planning and actions on public health.

## Funding

This work was supported by Sanofi Pasteur.

The funder of this collaborative study, participated in discussing the study’s findings and reviewing the manuscript. The authors analyzed and interpreted the data and submitted the manuscript for publication.

## Data Availability

All relevant data are within the manuscript and its Supporting Information files.

## Competing interests

The authors have declared that no competing interests exist.

## Data Availability

The databases used in the research will be made available in a repository at the Universidade Federal do Paraná, as soon as the publication of the article is confirmed.

## Notes

### Competing Interest Statement

The research received funding from SANOFI Pasteur, where the vaccine was developed and produced. The authors received grants to support the development of the research and declare a conflict of interest. However, this financial support did not influence the ethical commitments of the researchers, in the analysis and presentation of results, being faithful to what was found. Furthermore, the research was based on secondary data from government institutions and the researchers confirm that they will adhere to PLOS policies on sharing data and materials.

### Funding Statement

Yes

### Author Declarations

This study was approved by the Research Ethics Committee of the Federal University of Parana (Number 2,308,662). I confirm that all necessary patient/participant consent has been obtained and the appropriate institutional forms have been archived, and that any patient/participant/sample identifiers included were not known to anyone (e.g., hospital staff, patients or participants themselves) outside the research group so cannot be used to identify individuals.

